# Climate Change and Malaria: A Call for Robust Analytics

**DOI:** 10.1101/2024.09.16.24313623

**Authors:** Daniel J Laydon, David L Smith, Kaustubh Chakradeo, Mark P Khurana, Jaffer Okiring, David A Duchene, Samir Bhatt

## Abstract

Mosquitoes display marked sensitivity to weather, particularly temperature and precipitation. Therefore, climate change is expected to profoundly affect malaria epidemiology in its transmission, spatiotemporal distribution and consequent disease burden. However, malaria transmission is also complicated by other factors (e.g. urbanization, socioeconomic development, genetics, drug resistance) which together constitute a highly complex, dynamical system, where the influence of any single factor can be masked by others.

In this study, we aim to re-evaluate the evidence underlying the widespread belief that climate change will increase worldwide malaria transmission. We review two broad types of study that have contributed to this evidence-base: i) studies that project changes in transmission due to inferred relationships between environment and mosquito entomology, and ii) regression-based studies that look for associations between environmental variables and malaria prevalence. We then employ a simple statistical model to show that environmental variables alone do not account for the observed spatiotemporal variation in malaria prevalence.

Our review raises concerns regarding the relationship between climate change and malaria. We find that, while climate change’s effect on malaria is highly plausible, empirical evidence is much less certain. Future research on climate change and malaria must become integrated into malaria control programs, and understood in context as one factor among many. Our work outlines gaps in modelling that we believe are priorities for future research.

## Introduction

Malaria epidemiology, transmission, ecology and control are complex. The distribution of malaria, and its transmission intensity and seasonality have been shaped by a range of factors: climate [1–3], mosquito ecology and biogeography [4], malaria control [5], economic development [6], human genetics [7,8], and human history [9]. Since the start of the 20^th^ century, malaria incidence has declined over time, albeit unevenly [10, 11]. This geographical contraction [3] is primarily attributed to economic development and improved control strategies, such as healthcare strengthening and medical interventions. Conversely, factors such as drug resistance [12], changes in first-line therapies, pauses in malaria control, and the evolution of insecticide resistance continue to drive resurgence [13–15]. Aggregating these factors to identify individual contributions to incidence remains a significant analytical challenge.

Because malaria parasites are transmitted by mosquitoes, the interactions between weather and mosquito behavior have been of longstanding interest [1, 16, 17]. Here, we evaluate the evidence that has shaped current understanding and policy frameworks regarding climate change and malaria. We focus on the sources of uncertainty underlying this evidence, explaining that statistical studies suggest environmental variables primarily define a population at risk, rather than the actual current transmission intensity. Consequently, the influence of climate change on future malaria transmission appears highly uncertain.

Recent high-level policy frameworks illustrate the need for this re-evaluation. For example, the 2023 WHO World Malaria Report [18] includes a dedicated chapter on climate change, identifying it as a primary threat to global gains. Similarly, the IPCC Sixth Assessment Report (AR6) [19] notes with high confidence that climate change has influenced the distribution of vector-borne diseases, while simultaneously acknowledging the significant analytical challenge of disentangling these effects from concurrent trends in socioeconomic development and disease control.

We consider two kinds of studies that have examined the relationship between climate change and malaria. First, we consider studies of potential malaria transmission, which project changes in malaria transmission based on the link between environmental variables and either mosquito behavior or mosquito ecology. These studies model malaria transmission using either the basic reproductive number, *R*_0_, vector capacity [20, 21], or mathematical models of malaria transmission dynamics. In these studies, projections of malaria transmission under various climate change scenarios are based on observations of mosquitoes raised in a laboratory, in a semi-field environment, or in carefully controlled settings.

Second, we consider regression-based studies that look for associations between environmental variables and malaria prevalence. These studies have generally relied on large data sets curated by the Malaria Atlas Project [5]. We then use a simple regression to examine how environmental variables explain observed variation in malaria incidence.

We argue that methodological limitations, such as the omission of covariates for treatment failure and drug resistance, undermine the evidence for a dominant, independent link between climate change and malaria. To improve predictive utility, research on climate change should be integrated into broader malaria control and eradication goals [22].

## Potential Transmission

### Classical Malaria Dynamics

Many studies evaluating climate change and malaria are based on classical transmission dynamics, specifically Macdonald’s formula for the basic reproductive number, *R*_0_. Within this framework, *R*_0_ describes the number of human malaria cases arising from a single human case [20, 21, 23, 24].

Originally developed in the 1950s to synthesize early malaria epidemiology, [20, 25–32], Macdonald’s formula was designed to determine the critical mosquito density required to sustain transmission.

Macdonald used this mathematical model to weight the relative importance of specific parameters [20], particularly the sensitive role of mosquito survival [27]. The formula would serve as a threshold criterion: transmission would be sustained if *R*_0_ > 1. Therefore, to eliminate malaria, mosquito population density must reduce by at least a factor of *R*_0_, other parameters notwithstanding.

To develop theory for vector control, Garrett-Jones isolated the purely entomological parameters in the *R*_0_ formula and called the new formula “vectorial capacity” (VC, see Box 1) [33]. Ignoring differences among vector species in their ability to host parasites (since referred to as “vector competence”), VC describes a daily reproductive number: the number of infective bites that arise from all mosquitoes that blood-feed on a single, perfectly infectious human per day.

After early field trials exposed limitations of the Ross-Macdonald model [34], malaria models were extended to consider: immunity [35, 36]; treatment with antimalarial drugs and chemoprotection [37]; heterogeneous transmission [38–40], and mosquito ecology [36]. Such models have become embedded in comprehensive individual-based simulation models [41, 42], and used to guide malaria policies, including integrated malaria control [**?**].

Early studies on climate change and malaria employed computer simulation models, based on either simple extensions of the Ross-Macdonald model, or based on vectorial capacity (Fig 1, Box 1) [24, 43–46], which itself is a source of uncertainty [47]. A notable limitation of these earlier models was the absence malaria control interventions [41]. Consequently, while these classical models provide a vital mechanistic understanding of how temperature affects mosquito bionomics, they often lack the covariates necessary to account for the changing malaria patterns of the 20^th^ century, such as intervention coverage and drug resistance [41].

**Fig 1.**
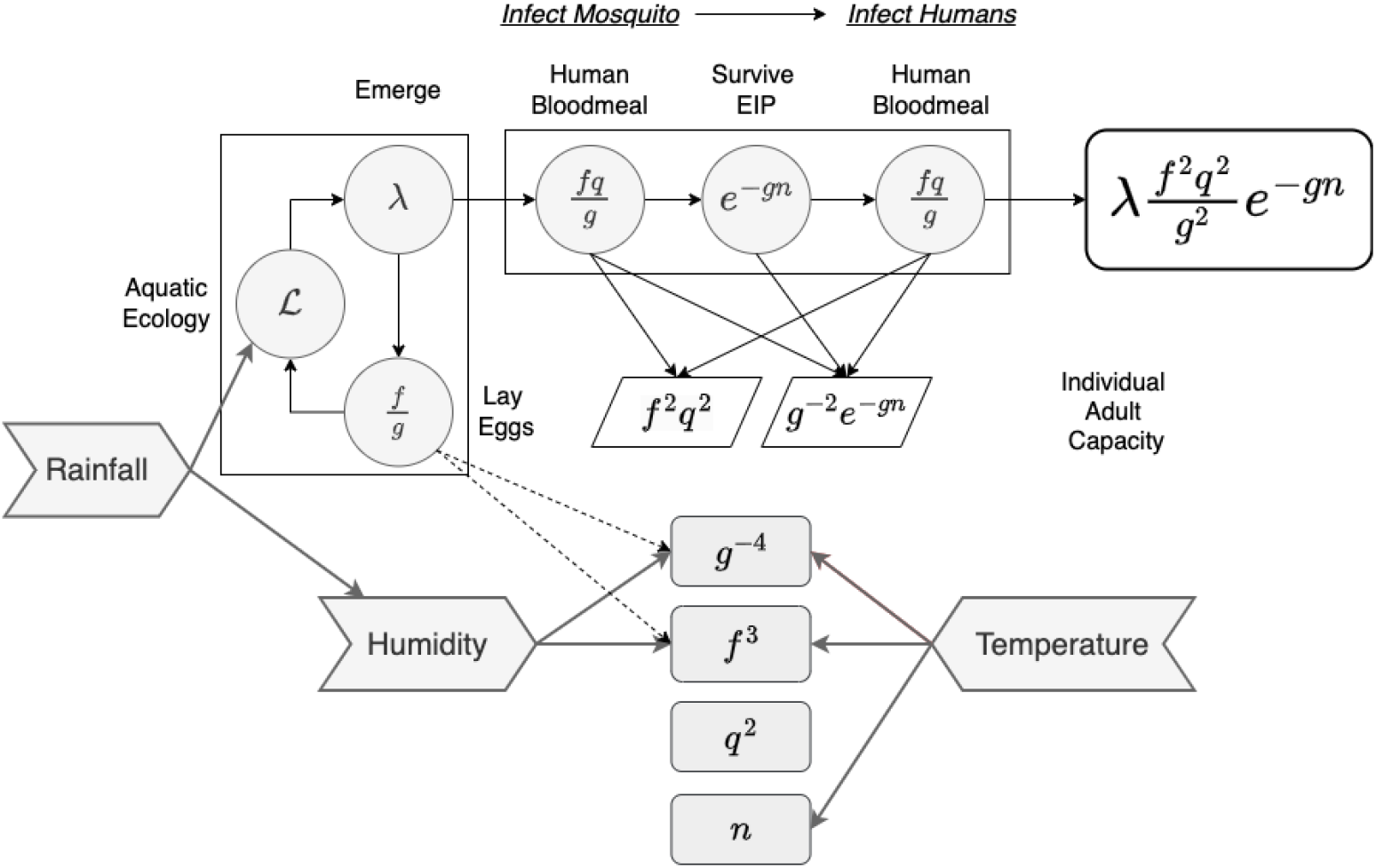
A diagram of 1) vectorial capacity as a summary of transmission potential (see Box #1) involving two parts: the emergence rate of mosquitoes, per human (*λ*); and the capacity of each individual mosquito to transmit parasites (*f* ^2^*q*^2^*e*^*−gn*^*/g*^2^.), where *f* is the blood feeding rate, *q* is the fraction of human blood meals among all blood meals, and *g* is the instantaneous death rate 2) Some of the likely effects of weather; and 3) a ranking of parameters by the number of ways they affect transmission. The box around mosquito aquatic ecology (ℒ), including egg laying by adults and emergence, indicates an important source of variability in malaria transmission that is also affected by weather in ways that often depend on the local context.

### Potential Transmission by Adult Mosquitoes

If a given parameter is related to temperature over time, *T* (*t*), it can be expressed as a function of both time and temperature1. For example, the mosquito death rate, *g*, can be written as *g*(*T* (*t*))2. Within this framework, a change in temperature results in a change to the reproduction number, *R*_0_, by a factor *Z*_*g*_:

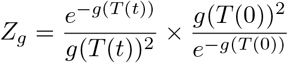

*Z*_*g*_ represents the effect size on potential transmission associated with temperature-driven changes in mosquito survival. Using the formula for vectorial capacity, it is possible to compute additional changes in potential transmission associated with feeding rates or the extrinsic incubation period (EIP), defined as the mean time required for parasites to undergo development within the mosquito before becoming infectious to humans. The total effect size on potential transmission by adult mosquitoes is the product of changes in each of these bionomic parameters.

Ideally, a total effect size should account for all changes caused by relevant environmental variables. An advantage of these metrics is their clarity in communication; e.g. an effect size of 1.5 can be reported as a 50% projected increase in potential transmission. These estimated effects are derived from field studies or controlled experiments measuring the impact of temperature and humidity on adult mosquito behavior, demography, and parasite development rates [48].

When Macdonald wrote his synthesis in 1952, dozens of studies had already measured the EIP in relation to temperature [27, 49]. More recent studies have examined the relationship between the EIP and temperature in *An. gambiae* and *An. stephensi* in high detail [50].

It is also useful to consider temperature through its interaction with relative humidity [51], which describes how much moisture the air holds relative to its maximum. The hotter the temperature, the more humidity the air can hold. There are strong associations between relative humidity and malaria transmission, and relative humidity also affects parasite and pathogen development within mosquitoes. Furthermore, relative humidity affects thermal performance curves of both mosquitoes and pathogens, leading to complex variation in the thermal optimum, limits, and operative range. Later studies addressed the impact of other environmental variables on vectorial capacity [52].

These studies largely agree on core messages [53]: mosquito daily survival and blood feeding rates, as well as the EIP, reach their optimum between 25 and 30 degrees, heavily dependent on relative humidity.

These biological relationships are well-evidenced, and the primary challenge is now how to integrate these laboratory-derived effect sizes into the broader, non-linear context of actual disease prevalence in the field.

### Climate and Mosquito Populations

Since standing water serves as mosquito habitat, rainfall creates opportunities for exponential mosquito population growth. However, changing rainfall patterns are highly unpredictable; short-term rainfall is inherently volatile, and long-term climate projections for precipitation carry even greater uncertainty. Furthermore, the effects of rainfall on mosquito ecology are locally idiosyncratic, mediated by hydrology, terrestrial ecology, and numerous other factors. Consequently, the relationship between rainfall and transmission may be specific to each locality, where a single rain event can drive diverging patterns for local vector species populations [54].

The influence of rainfall on malaria transmission relates fundamentally to the availability and quality of mosquito habitats. Any concavity filled by rainfall or subterranean water flows can become a habitat for immature mosquitoes. However, increased precipitation does not necessarily lead to more habitat or higher mosquito densities. First, rainfall is mediated by complex local hydrology. Second, its impact is affected by temporal distribution—the magnitude of events and the duration between them. Third, aquatic mosquito populations are affected by many biotic interactions, including resource competition.

While temperature tends to vary smoothly across landscapes, rainfall exhibits high spatial variation [55]. For instance, increased egg laying in a crowded habitat could delay mosquito development, and lower the number of adults emerging. And while rainfall can increase the number and size of breeding sites, excess rainfall can wash them out [56].

Additionally, rainfall is a non-equilibrium relaxation process that is scale free, and best described by a simple power law, characterizing the density and occurrence of rain events, as well as drought periods. That is, rainfall events can be of enormous size in a very short period, followed by a prolonged drought, exhibiting complex and unpredictable fluctuations over time. Incidentally, earthquakes are another example of a non-equilibrium relaxation process [57].

In the context of climate change, warmer oceans increase evaporation, intensifying precipitation and storm systems. While the resulting increased rainfall is expected to increase standing water and therefore mosquito habitats, it is also expected to increase both floods and droughts, both of which can paradoxically reduce malaria transmission.

These dynamics suggest that average rainfall is often a misleading indicator of true transmission risk. Therefore, identifying the effects of climate change is even more challenging, and requires moving beyond macroscopic scale predictions toward context-dependent models that account for local resource availability and habitat stability

### Sensitivity, Variability, and Uncertainty

A common mathematical approach to examining the potential effects of climate change on mosquitoes is to evaluate the sensitivity of transmission to specific biological parameters [58]. While many studies have emphasized the importance of adult mosquito survival and blood feeding, an examination of the data suggests that the majority of variability in transmission is actually driven by mosquito ecology [59]. The primary empirical data for these assessments come from studies that estimate the entomological inoculation rate (EIR), which is computed as the product of the human biting rate (HR) and the sporozoite rate (SR).

The annual EIR can range from practically zero to more than a thousand infectious bites per person, per year [60]. Crucially, the overwhelming majority of this variability is attributed to differences in the HR rather than the SR. This implies that transmission scales primarily with adult mosquito population density, which is governed by complex aquatic ecology and habitat availability, rather than just the temperature-dependent speed of parasite development within the mosquito.

While it is therefore reasonable to suggest that transmission should scale linearly with mosquito density, mosquito ecology is inherently non-linear. A single adult female can produce thousands of offspring over a few days, leading to explosive, non-linear bouts of transmission that are difficult to capture in models that rely on average temperature or rainfall values. This highlights a research gap: if the drivers of mosquito density are more influential than the drivers of parasite development speed, then climate models that ignore local ecological constraints and larval habitat dynamics may significantly over- or under-estimate the impact of warming.

### Complexity, Scaling, and Malaria Metrics

To understand the effects of changes in potential transmission, mathematical models are required to determine how malaria in humans responds to shifts in transmission intensity. Such models highlight non-linearities and complexities in the relationships between infectious mosquito bites, parasite infections and disease burden.

The relationship between the EIR and the average parasite rate (PR) is strongly non-linear [61, 62]. While the EIR measures exposure, the associated force of infection (FoI) and malaria incidence are also non-linear and vary significantly by age [63, 64]. Malaria immunity develops over time with exposure, meaning disease is often concentrated in young children. Changes in transmission intensity are expected to shift this burden to older age groups, but the overall change in burden is not a simple linear response to increased mosquito density.

These non-linearities create substantial uncertainty regarding how one parameter affects another [64] (e.g. doubling the VC or EIR may not double mortality), and make it inherently difficult to produce credible projections about the changing burden of malaria, even in idealized scenarios where expected changes in transmission are precisely quantified. This complexity suggests that focusing on climate as a sole driver may overlook the stabilizing effects of immunity and the heterogeneous response of different age groups to shifting transmission levels.

### Thresholds, Importations and Heterogeneity

Models and Macdonald’s threshold condition have led to concerns that *R*_0_ may increase above one due to climate change1. While Macdonald’s formula for *R*_0_ was intended to describe a threshold condition for the establishment of endemic transmission, the predicted effects of crossing such a threshold are often dulled by malaria connectivity.

Malaria transmission can be sustained by importation in mobile human populations. In models with spatial dynamics, threshold conditions are modified by heterogeneity, and transmission is dispersed widely by the movement of humans and mosquitoes [65].

In most places where *R*_0_ *<* 1, malaria is sustained through importation; therefore, crossing a theoretical threshold may not lead to a qualitative change in local epidemiology. Three important factors modifying these conditions are the heterogeneous spatial distribution of mosquitoes, the heterogeneous distribution of humans, and the temporal variability of transmission potential. Consider a simple conceptual model for a time-varying reproduction number *R*_0_(*t*) that is piecewise constant (i.e. a step function), where seasonal endemic transmission is characterized by periods where *R*_0_(*t*) > 1, and other periods where *R*_0_(*t*) ≤ 1. However, empirical malaria data is highly heterogeneous [59], and estimates of reproduction numbers exhibit roughness in their functional form over time [66] and space [3, 67].

Parasite populations are connected by movement of infected humans and mosquitoes [65]. So while Macdonald’s formula *R*_0_ > 1 remains a vital conceptual tool, it is best regarded as an informative pseudo-threshold in the context of connected populations [65, 68–71].

### Predictions

Sub-Saharan Africa carries the majority of worldwide malaria burden, and so climate change’s effects here are of particular interest. Despite uncertainty about the shape of *R*_0_(*T*), several lines of evidence suggest that the marginal effects of warming are likely to be small in many regions.

First, across much of Africa, temperatures are near the thermal optimum where minor changes have negligible effects on potential transmission. Second, much of Africa is already above the optimum, suggesting that an increase in temperature could likely decrease transmission in those areas. Third, the predicted effect sizes of climate on potential transmission are much smaller than the potential reductions in transmission that can be achieved through vector control [3]. Fourth, the greatest changes in malaria transmission are likely to come through changes in rainfall [2].

The recent Malaria Altas report https://malariaatlas.org/project-resources/climate-change/ on climate change is consistent with the above conclusions. It estimates that there will be an additional 550,000 malaria deaths by 2050, 90% of which are due to extreme weather events. Given that estimated annual malaria mortality is 500,000 deaths, climate change is expected to add approximately only 1/25 of annual burden.

## Regression

A common approach to climate change and malaria has come through regression analyses [3, 5, 72, 73], which have been used to interpolate data across space and time to understand the drivers of change [5].

Here, we perform a simple regression analysis using detailed satellite imagery, to collect data on temperature and precipitation. These data are matched to the location and time (month) of malaria observations, which we denote as *y*. These observations are adjusted for age (2 - 10 years) and diagnostic type [74]. Our statistical model is expressed as:

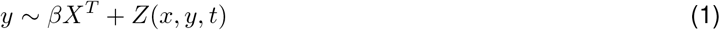

In this equation, *y* represents the *Pf* PR (*Plasmodium falciparum* parasite rate)), *X* is a vector of environmental covariates, *β* is a vector of coefficients, and *Z* is a zero-mean Gaussian process with a space-time covariance function denoting the residuals.

Intuitively, this model attempts to explain PfPR as a function of environmental covariates *X*. If the predictive power of this function is high, an argument can be made for a direct relationship between climate variables and malaria prevalence. The *Z* term accounts for residual patterns caused by unobserved factors, such as nutrition, mosquito ecology, or human mobility. We fit this model using Approximate Bayesian inference (the Laplace approximation [75]) such that the resultant model balances over- and under-fitting.

For a simple illustration, we consider two major climate factors: temperature suitability and average rainfall. Temperature suitability [76, 77] is a dynamic biological mathematical model that incorporates temperature dependency in the malaria transmission cycle, and then uses satellite data on temperature to estimate a suitability index. Rainfall is measured using CHIRPS (Climate Hazards Group InfraRed Precipitation with Station data), which estimates the average rainfall per month from rain gauge and satellite observations. It is then possible to match this temperature suitability index to the month and year of malaria observations, and to match rainfall to the month (averaged over years) to account for minor aspects of seasonality.

Outliers such as heatwaves, droughts and floods are not adequately captured using these data, but major variations in the spatial and temporal distribution of the environment factors relevant to the mosquito are. Using the large Malaria Atlas Project dataset on malaria parasite rate surveys, where each data point is a sample of the number of parasite positive individuals out of the total, it is possible to match temperature suitability to the specific latitude, longitude, rainfall, month and year (2001-2022) to the specific latitude, longitude, month, with years averaged due to data paucity. For consistency, data on parasite rates are adjusted for age [78] and diagnostic time [74].

Malaria parasite rate data are proportions, thus bounded between zero to one. To simplify regression, we transformed these data via the empirical logit into a Gaussian scale. Once again, we call these observations *y*. Consider three simple models, explaining parasite rate observations by: (i) a constant model *y*_*x,y,t*_ ∼ *I*; (ii) a linear model with temperature suitability and rainfall *y* ∼ *I* + *β*^*T*^ *X*_*x,y,t*_, where *X*_*x,y,t*_ is the temperature suitability index and rainfall at the matched locations and times of the malaria parasite rate observations; and (iii) a Gaussian process model *y* ∼ *I* + *β*^*T*^ *X*_*x,y,t*_ + *Z*(*x, y, t*), where again *Z* is a space-time random field that captures structure in the data. We evaluate model performance by computing the mean absolute percentage error and the correlation on the original untransformed parasite rate scale.

The results are revealing: the first model fitting a simple constant intercept has a correlation of zero (with a mean absolute error of 17%). The second, linear model with temperature suitability does explain variation with a mean absolute error of 16.5% and a correlation of 0.2. The third Gaussian process model with temperature suitability yields a mean absolute error of 9% and a correlation of 0.8. This difference is substantial, and while this example is simplistic, and by no means rigorous, it reveals that the bulk of the spatial distribution of malaria and its change in time over the past two decades is negligibly explained by temperature and its biological effect on the mosquito.

The substantial discrepancy in predictive power between the linear climate model and the Gaussian process identifies a quantifiable “information gap” in current climate-centric analyses. This gap suggests that the variance in malaria prevalence is largely driven by unobserved non-climatic factors, e.g. health system strength and socioeconomic development, which are often omitted from stand-alone climate projections. Filling this gap is essential for creating models that are operationally useful for public health planning rather than merely illustrative of environmental suitability

Figure 2 shows the predictions for the linear and Gaussian process model (i.e. the second and third models) alongside the raw data. We see that the model with just temperature suitability and rainfall is unable to capture the large variations in parasite rate, and creates predictions within a narrow band (Figure 2 top right) and the fine grained spatial variation only predicts a limited variation in *Pf Pr* (Figure 2 bottom left). In contrast, the Gaussian process (Figure 2 bottom right) is an excellent fit to the data, both in terms of spatial pattern but also in predicting the full range of variation in *Pf Pr*. These results reinforce that, while climate plays a pivotal role in defining the population at risk, simple relationships are not the primary driver in the dynamic changes of infection.

**Fig 2.**
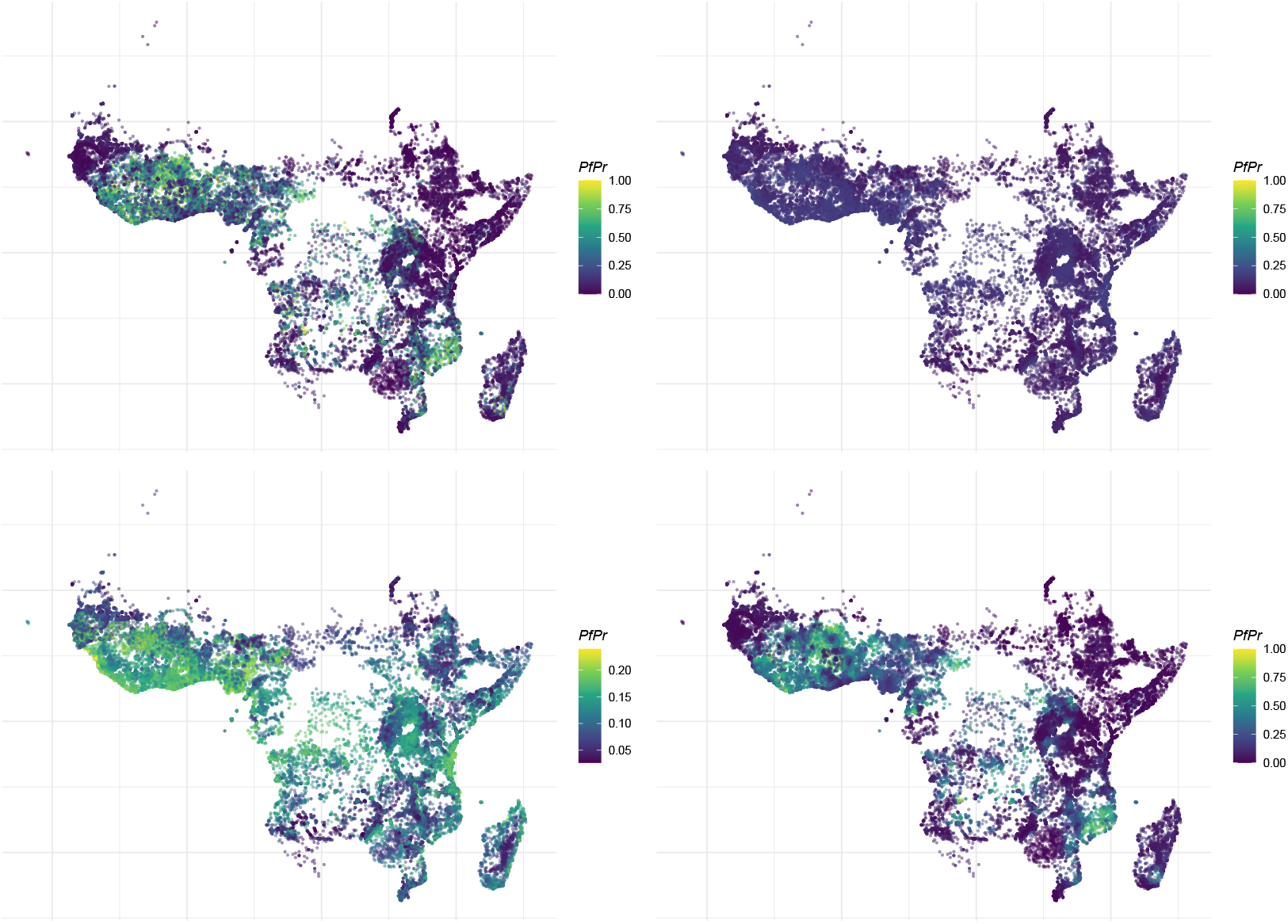
*P. falciparum* prevalence or parasite rate (*Pf* PR_2*−*10_) as a function of environmental covariates. (top left) *Pf* PR from the Malaria Atlas Project Database. (top right) Linear model with temperature and rainfall with the colour scale ranging from 0-1 (bottom left) Linear model with temperature and rainfall with a restricted colour scale to show variation (bottom right) Gaussian process process with linear mean function of temperature and rainfall

Model code is available at https://github.com/dlaydon/MalariaClimateRegression.

## Gaps in modelling

While substantial efforts have been made to model malaria dynamics over time [41], important gaps remain for future research to address. In most settings, malaria transmission is best understood as a shifting baseline that has been modified by intensive malaria control. While many studies have examined climate effects on baseline malaria, and others have focused on vector control, few have rigorously examined the interactions between the two.

In future climate and malaria research, it is important to consider the role of exogenous variables affecting the baseline. Yearly fluctuations in temperature and rainfall are often greater than the projected changes in their average values. Therefore, further studies of the factors driving seasonality are likely to be especially informative on the likely impact of climate change [79]. Although factors such as changing housing quality, demography, and the evolution of drug and insecticide resistance are easy to list, including them in longitudinal analyses is often hampered by a lack of consistently available data.

Since 2000, the malaria landscape has been profoundly altered by the mass distribution of long-lasting insecticide-treated nets and widespread access to artemisinin-based combination therapies (ACTs) [5].

Understanding the effects of these interventions is currently hindered by a lack of granular knowledge regarding local vector species mixes, ecology, and resistance patterns. Furthermore, data on socioeconomic factors and local environmental conditions are necessary to understand how topography influences transmission within any given region. Addressing these data gaps will allow for the greater integration and cooperation of climate work with other strands of malaria research and policy.

### Integrated surveillance

To foster greater cooperation between climate work and malaria policy, we argue that research should move toward “integrated surveillance”. Rather than producing stand-alone long-term risk projections, climate data should be utilized within multi-factorial frameworks to optimize the timing and location of vector control interventions. This requires using shared datasets to model environmental suitability and control efficacy simultaneously, ensuring that climate science serves the central task of burden reduction.

### Drug resistance

Drug resistance profoundly influences malaria independently of climate change. The widespread use of antimalarial drugs has led to strong selection for resistant parasite strains, which can arise from relatively modest molecular changes. For example, just four amino acid substitutions in the *pfcrt* (*P. falciparum* chloroquine resistance transporter) gene confer resistance to chloroquine-based drugs [80].

While there is a hypothetical link between climate and resistance—where climatic conditions might mediate the logistics of drug administration [81] or disproportionately benefit the emergence of resistant strains—the biological mechanisms remain largely undescribed. Moving forward, genomic surveillance can play a vital role in optimizing drug administration by mapping the spread of resistant strains. Integrating such genetic data with climate models would provide a more holistic view of the challenges facing malaria eradication [82].

## Conclusion

Our work highlights significant sources of uncertainty regarding climate change and malaria research, specifically concerning the complexity of the processes involved. Potential effect sizes due to temperature are often masked or mimicked by other factors, including vector control, anti-malarial drug use, and socioeconomic development. Given this complexity, climate-related risks must be evaluated within multi-factorial studies that provide accurate assessments of causation. Our analysis suggests that the risks posed by climate change on malaria incidence may be modest relative to other drivers when accounting for observed spatiotemporal variability.

In many settings across sub-Saharan Africa, current temperatures are near the optimum for transmission; therefore, future warming is as likely to reduce transmission as it is to increase it. Empirical data emphasize that rainfall, and its interaction with temperature and humidity, remains a critical but highly context-dependent driver. Methodological challenges, such as the failure to consider the evolution of chloroquine resistance during the 1990s, have historically undermined the validity of longitudinal climate-malaria studies. While climate change will likely cause spatially heterogeneous shifts in transmission, effective ways of managing these effects are already available.

To summarise, we have attempted to re-evaluated the evidence for climate-driven shifts in malaria epidemiology through reviewing relevant literature and via simple statistical analysis. While biological models confirm that temperature and rainfall define environmental suitability, our regression results demonstrate that these variables alone explain only a relatively small amount of the dynamic shifts in infection prevalence observed over the past two decades. The high performance of the Gaussian process model, relative to the linear climate-only model, suggests that malaria prevalence is governed more by unobserved socioeconomic and public health factors than by climatic trends in isolation.

Ultimately, these findings indicate that the influence of climate change on future malaria transmission is highly uncertain, despite the fact that it remains highly plausible. We argue that research would best be directed away from isolated sensitivity analyses toward integrated, multi-factorial frameworks that explicitly account for the shifting trends observed over the last century, such as changing intervention coverage and the evolution of drug resistance. By situating climate risk within the broader context of malaria control and economic development, researchers can provide the nuanced insights required to protect vulnerable populations in a changing environment.

### Box 1: Vectorial Capacity

Macdonald’s *R*_0_ formula was based on the sporozoite rate (SR) and the human biting rate (HR) [20, 83], which is now called the entomological inoculation rate (EIR).

Vectorial capacity (VC) includes three parameters: the blood feeding rate (*f*); the fraction of all blood meals that are human blood meals (*q*); and instantaneous death rate (*g*). Together, these terms describe the expected number of human bloodmeals a mosquito would take over its lifetime (*S* = *fq/g*). A single parameter describes parasites in mosquitoes, called the extrinsic incubation period (EIP, *n* days), defined as the number of days required for malaria parasites to develop. To transmit, a mosquito must survive through the EIP (with probability *P* = *e*^*−gn*^). The formula for VC includes one parameter describing mosquito ecology: the emergence rate of mosquitoes from aquatic habitats, per human (*λ*). These parameters are combined into a formula for vectorial capacity.

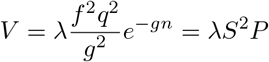

The right hand side denotes parasite transmission by mosquitoes: after emerging (*λ*), a mosquito must blood feed on a human to become infected (*S*), then survive through the EIP (with probability *P*); and then bite other humans to transmit (*S*). This is equivalent to Macdonald’s formula, after a change in notation [59]. The formula has been used to understand parameters that could have the greatest influence on transmission [47, 83].

### Box 2: Mosquito Ecology

Mosquitoes have seven distinct life stages: eggs, four larval instars, pupae, and adults (Figure 3). Adults lay eggs in aquatic habitats. After hatching and developing in water through pupation, adults emerge as adults that mate and sugar feed. Female mosquitoes (but not males) also blood feed; the protein and nutrients in blood are used to make eggs. It is the cycle of blood feeding, egg laying, and sugar feeding by adult females that is of greatest interest sets the stage for mosquito ecology and malaria parasite transmission. Mosquitoes, like most insects, are poikilothermic – their internal temperature depends on the surrounding environment. Mosquito activities and many of the resources they require to complete their life cycle are also affected by weather and climate, including vertebrate animals for blood, sugar, vegetation and resting habitats, and aquatic habitats.

**Fig 3.**
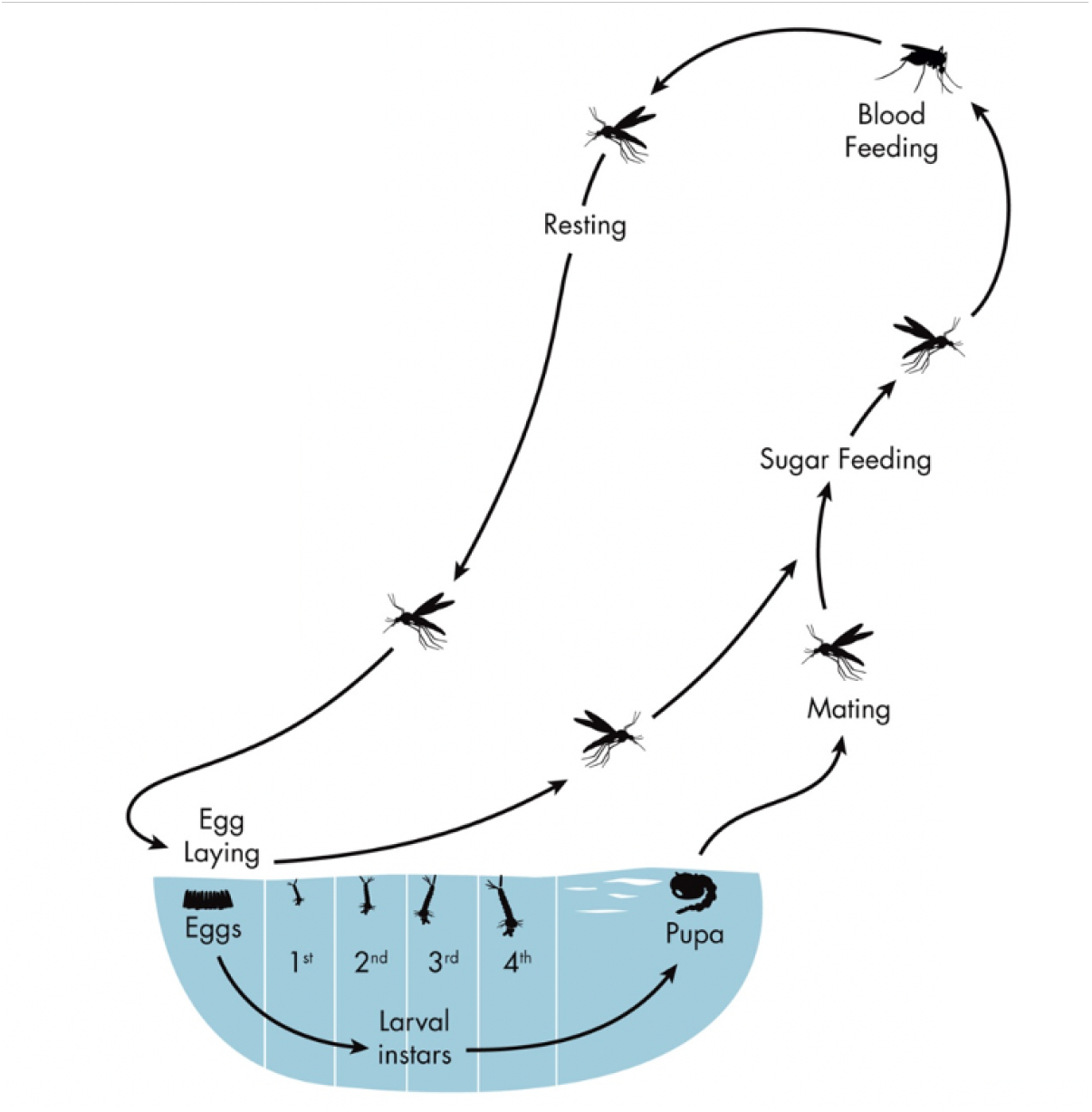
The mosquito life cycle includes immature aquatic stages and a volant adult. Female mosquitoes lay eggs in water bodies. Eggs hatch within a few days to months. Larvae live in water and develop into pupae in as few as 5 days. Pupae continue to live in water and develop into flying adult mosquitoes that leave the water in 2-3 days. Adult mosquitoes meanwhile fly in search of resources, including vertebrate hosts to blood feed, sugar sources for sugar, and aquatic habitats to lay eggs.

## Data Availability

All data in the present work are contained in the manuscript.

## Acknowledgments

DJL acknowledges funding from the Wellcome Trust for the Vaccine Impact Modelling Consortium (VIMC) Climate Change Research Programme (grant ID: 226727 Z 22 Z). DJL, SB acknowledge funding from the MRC Centre for Global Infectious Disease Analysis (reference MR/X020258/1), funded by the UK Medical Research Council (MRC). This UK funded award is carried out in the frame of the Global Health EDCTP3 Joint Undertaking. DLS and JO acknowledge funding from a grant from the Bill and Melinda Gates Foundation (INV 030600). DLS was also supported by a grant from the National Institute of Allergies and Infectious Diseases (R01 AI163398). SB is funded by the National Institute for Health and Care Research (NIHR) Health Protection Research Unit in Modelling and Health Economics, a partnership between the UK Health Security Agency, Imperial College London and LSHTM (grant code NIHR200908). Disclaimer: ‘The views expressed are those of the author(s) and not necessarily those of the NIHR, UK Health Security Agency or the Department of Health and Social Care.’. DAD acknowledges support from the Novo Nordisk Foundation via the Emerging Data Science Investigator award (NNF23OC0084647). SB acknowledges support from the Novo Nordisk Foundation via The Novo Nordisk Young Investigator Award (NNF20OC0059309), which also funds KC. SB acknowledges the Danish National Research Foundation (DNRF160) through the chair grant which also supports MPK. SB acknowledges support from The Eric and Wendy Schmidt Fund For Strategic Innovation via the Schmidt Polymath Award (G-22-63345).

